# Antimalarial Drug Resistance in *Plasmodium falciparum* from Tak Province, Thailand (1998-2001): A TaqMan Array Card Study

**DOI:** 10.1101/2024.05.29.24308164

**Authors:** Sasikanya Thaloengsok, Chaiyaporn Chaisatit, Piyaporn Saingam, Paphavee Lertsethtakarn, Michele Spring, Sriwichai Sabaithip, Suporn Pholwat, Jennifer L Guler, Eric Houpt, Brian Vesely

**Affiliations:** Walter Reed Army Institute of Medical Science-Armed Forces Research Institute of Medical Sciences (WRAIR-AFRIMS), Bangkok, Thailand; Division of Infectious Disease and International Health, Department of Medicine, University of Virginia, Charlottesville, VA, USA; University of Virginia, Charlottesville, VA, USA

**Keywords:** PfKelch13 Mutations, GMS, Plasmodium, TAC

## Abstract

Despite declining malaria cases in Thailand, surveillance in endemic areas is crucial. Retrospectively analyzing samples from Tak province, Thailand, we found prevalent drugresistant *Plasmodium falciparum*, particularly to mefloquine and sulfadoxine/pyrimethamine. Notably, mutations indicating resistance to artemisinin were detected at low frequencies, suggesting evolving resistance. These findings stress the need for continuous surveillance to guide control strategies and prevent outbreaks, even with decreasing cases, to sustain malaria elimination efforts.

---

Malaria remains a significant public health challenge in the Greater Mekong Subregion (GMS), historically serving as a hotspot for transmission by *Plasmodium falciparum* and *Plasmodium vivax*. Despite progress in reducing incidence over the past decade, transmission persists in border areas and remote regions with limited healthcare access (Sudathip et al., 2019). Artemisinin-based combination therapies (ACTs) have been crucial in treatment and control efforts, but artemisinin resistance threatens progress. PfKelch13 mutations are central to this resistance, altering the parasite’s response to artemisinin (Ashley et al., 2014). The WHO has identified thirteen validated PfKelch13 markers of resistance (World Health Organization, 2022).

The earliest documented PfKelch13 mutation linked to resistance dates to 2008, with subsequent high prevalence in *P. falciparum* isolates, especially in border regions and areas with extensive antimalarial drug use (Ménard et al., 2016). Surveillance systems, including the innovative *P. falciparum* TaqMan Array Cards (TACs), play a vital role in monitoring PfKelch13 mutations. TACs offer a high-throughput, cost-effective means of detecting genetic markers associated with *P. falciparum*, aiding large-scale surveillance and clinical trials (Pholwat et al., 2017).

Our study presents findings from a survey using TaqMan Array Cards (TACs) to assess drugresistant markers in Mae Sot, Tak, Thailand, from 1998, 1999, and 2001, offering insights into the prevalence and dynamics of artemisinin resistance-conferring PfKelch13 mutations, enhancing understanding of malaria and drug resistance in the GMS (Figure 1). A total of 808 archived blood samples were collected during malaria studies evaluating rapid diagnostic tests (RDTs) for P. *falciparum* in Tak Province, Thailand, conducted from 1998 to 2001. These samples were primarily assessed for both mono- and mixed *P. falciparum/P. Vivax* infections by smear and were subjected to analysis using TACs. Out of the total samples, 112 were excluded due to the absence of *Plasmodium* spp. or being positive only for *P. vivax*. The remaining 696 samples were confirmed to be positive for *Plasmodium* species, with *P. falciparum* being the most prevalent (Table 1).

**Table 1:** Distribution of *Plasmodium* spp. infections by TACs.

|  | N=30 | N=206 | N=460 |
| --- | --- | --- | --- |
| Year | 1998 | 1999 | 2001 |
| PF | 26 | 180 | 378 |
| PF+PV | 4 | 26 | 76 |
| PF+PM | 0 | 0 | 4 |
| PF+PV+PM | 0 | 0 | 1 |
| PF+PV+PO | 0 | 0 | 1 |
| Negative | 5 | 87 | 20 |

**Figure 1.**
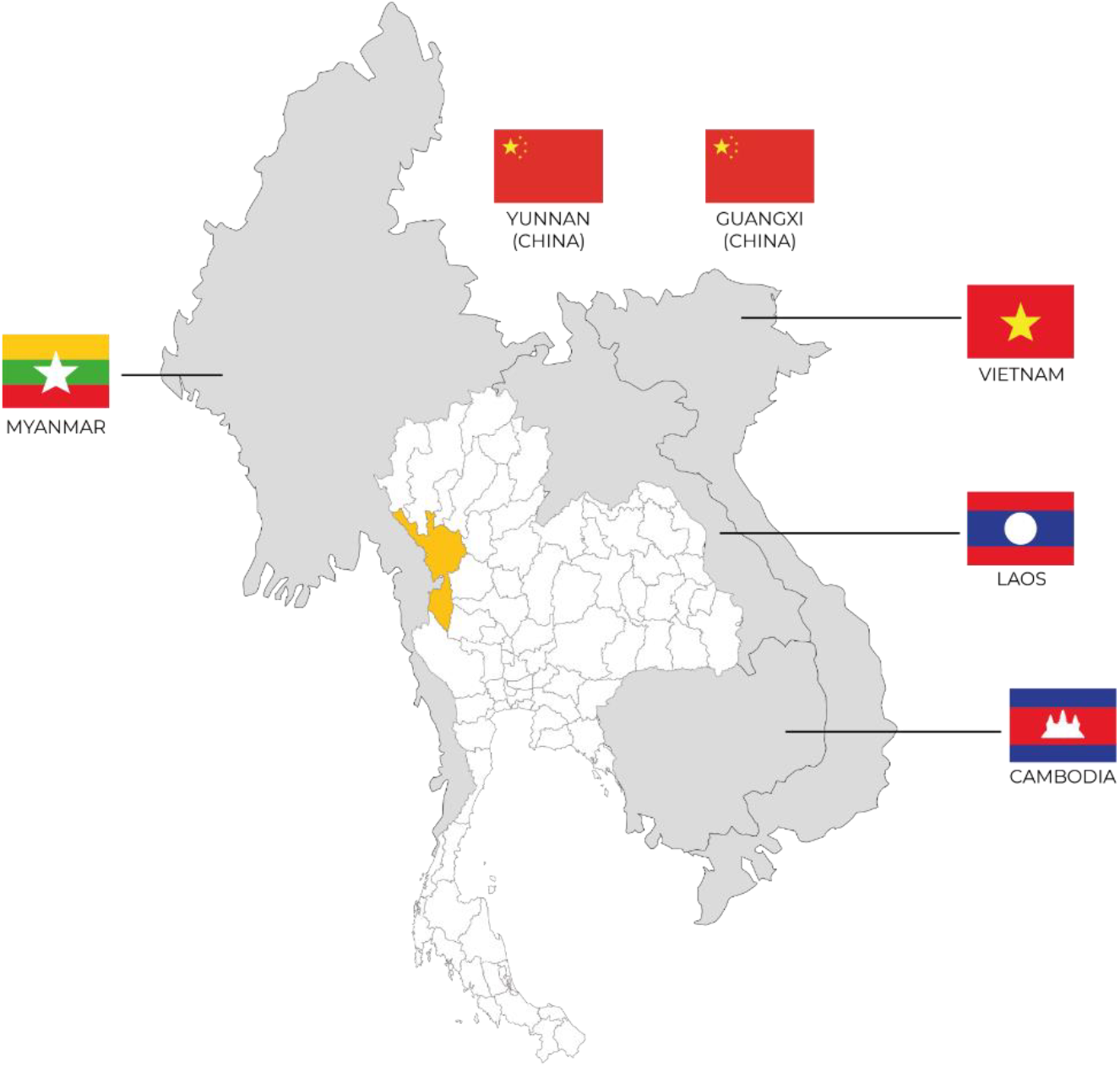
A map of the Greater Mekong Subregion (GMS) with the sample site highlighted in Mae Sot District, Tak Province, Thailand.

Using the *Plasmodium* TAC panel, mutations in several key genes associated with drug resistance and malaria biology were analyzed. These genes included PfKelch13, *P. falciparum* CQ-resistant transporter (Pfcrt), Pfdhfr, Pfdhps, cytochrome bc1, and Pfmdr1 *P. falciparum* genes (Table 2). The Pfcrt mutations at amino acids 72-76 reflect chloroquine resistance, selected for the TAC panel before the discovery of novel mutations’ role in piperaquine resistance (Muhamad et al., 2013).

**Table 2:**
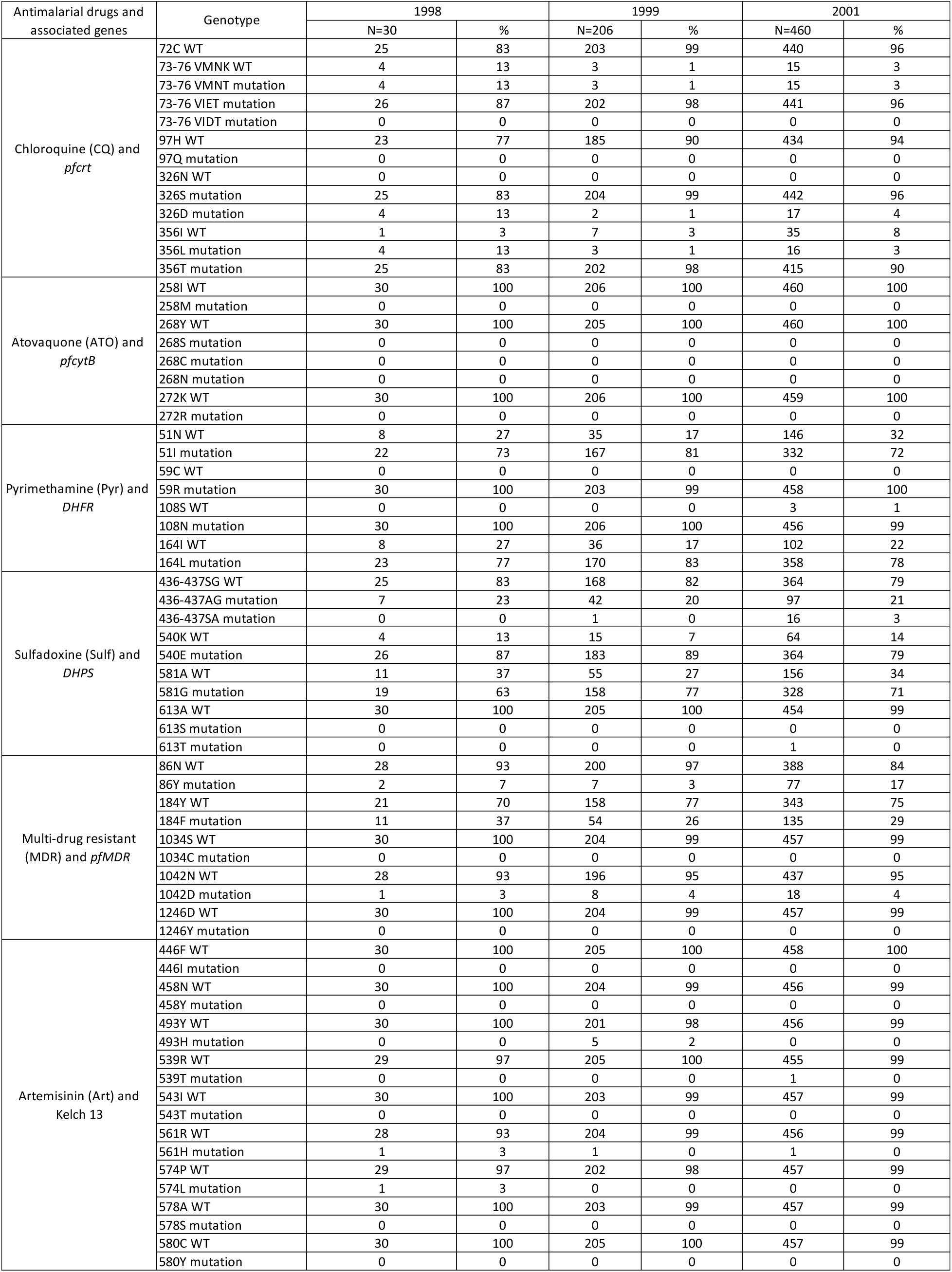
Summary of TAC results.

During the sample collection period, the first-line treatment for uncomplicated *P. falciparum* infection was mefloquine alone or in combination with sulfadoxine/pyrimethamine (S/P), later with artesunate. Mutations conferring mefloquine resistance were observed in Pfmdr1 at positions N86Y, Y184F, and N1042D. Additionally, mutations associated with S/P resistance, particularly in Pfdhfr and Pfdhps, were prevalent. Notably, a double mutation of Pfdhfr at positions 59R and 108N was highly prevalent (>99%), and multiple mutations were observed in Pfdhps, notably 540E and 581G. These mutations were indicative of a strong selection pressure imposed by the widespread use of these antimalarial drugs (Myo Thura Zaw et al., 2020).

Analysis of PfKelch13 mutations revealed the presence of mutations at positions P574L, Y493H, and R539T, all confirmed by the WHO to confer resistance to artemisinin (Table 3). These findings suggest the emergence of baseline resistance markers even in the absence of drug pressure, highlighting the adaptive nature of *P. falciparum* to changing drug regimens (Pholwat et al., 2017). Chloroquine ceased being used for *P. falciparum* due to high-level resistance, but it remained first-line for *P. vivax*. While the K to T mutation at position 76 of *pfcrt* is thought to be pivotal in resistance, the 72-76 CVIET combination, indicative of Indonesian lineage, and mutations in 326/356 have been at saturation for at least 15 years, present in 86-98% of samples. No mutations associated with atovaquone resistance were detected.

**Table 3.** PfKelch13 SNPS identified by TAC.

|  | 1998 |  | 1999 |  | 2001 |  |
| --- | --- | --- | --- | --- | --- | --- |
| Artemisinin (Art) and Kelch | N=30 | % | N=206 | % | N=460 | % |
| WT Kelch 446F | 30 | 100 | 205 | 100 | 458 | 100 |
| WT Kelch 458N | 30 | 100 | 204 | 99 | 456 | 99 |
| WT Kelch 493Y | 30 | 100 | 201 | 98 | 456 | 99 |
| mutation Kelch 493H | 0 | 0 | 5 | 3 | 0 | 0 |
| WT Kelch539R | 29 | 97 | 205 | 100 | 455 | 99 |
| mutation Kelch 539T | 0 | 0 | 0 | 0 | 1 | 0 |
| WT Kelch 543I | 30 | 100 | 203 | 99 | 457 | 99 |
| WT Kelch 561R | 28 | 93 | 204 | 99 | 456 | 99 |
| mutation Kelch 561H | 1 | 3 | 1 | 0 | 1 | 0 |
| WT Kelch 574P | 29 | 97 | 202 | 98 | 457 | 99 |
| mutation Kelch 574L | 1 | 3 | 0 | 0 | 0 | 0 |
| WT Kelch578A | 30 | 100 | 203 | 99 | 457 | 99 |
| WT Kelch 580C | 30 | 100 | 205 | 100 | 457 | 99 |
| mutation Kelch 580Y | 0 | 0 | 0 | 0 | 0 | 0 |

Artemisinin resistance in *P. falciparum* has presented a significant challenge to malaria control efforts in the GMS. Monitoring mutations in the PfKelch13 gene, some strongly associated with artemisinin resistance, has aided national malaria programs in adjusting first-line therapies and increasing control efforts to inhibit transmission (Ménard et al., 2016). With PfKelch13 prevalence increasing in East Africa, understanding the emergence and spread of PfKelch13 mutations may assist in the prediction and interruption of drug failures there.

The study contributes to understanding artemisinin resistance dynamics by detecting the P574L, Y493H, and R539T mutations in samples collected from Tak, Thailand, between 1998 and 2001. These mutations were previously reported in Thailand in 2009, highlighting the persistence and spread of resistance alleles over time. The findings underscore the early emergence of resistance in the GMS, consistent with WHO assessments (Ashley et al., 2014). Comparisons with earlier and subsequent studies could provide insights into mutation trends over a decade. The study also ties into related research, emphasizing the importance of ongoing surveillance and intervention efforts to combat anti-malarial resistance. Tools such as TACs offer valuable insights into resistance dynamics, aiding in the development of targeted interventions to address the evolving challenges posed by malaria.

The importance of continuous surveillance to monitor resistance profiles and inform malaria control strategies cannot be overstated. Even amid declining malaria case numbers, vigilance is crucial to prevent outbreaks and safeguard elimination efforts. The presence of drug-resistant strains, particularly those resistant to artemisinin, threatens to undermine the progress made in malaria control. The adaptive nature of *P. falciparum*, demonstrated by the observed mutations, highlights the need for adaptable and robust malaria management programs. As resistance markers continue to evolve, the use of advanced molecular techniques, such as TACs, becomes indispensable in providing timely and accurate data to guide public health interventions.

This study of archived blood samples from Tak Province, Thailand, using TaqMan Array Cards has provided valuable insights into the prevalence and types of drug-resistant mutations in *P. falciparum* and to our knowledge this is the oldest reported PfKelch13 mutations found in the GMS. The identification of key mutations associated with resistance to chloroquine, mefloquine, sulfadoxine/pyrimethamine, and artemisinin underscores the complex and evolving challenge of malaria control. These findings highlight the necessity for ongoing surveillance and adaptive public health strategies to combat the persistent threat of drugresistant malaria. Advanced molecular tools and comprehensive genetic analysis are crucial in informing and guiding these efforts, ensuring that control measures are both timely and effective. The fight against malaria requires constant vigilance, innovation, and a commitment to understanding the ever-changing landscape of antimalarial resistance.

## Data Availability

All data produced in the present study are available upon reasonable request to the authors

## Disclaimer

Material has been reviewed by the Walter Reed Army Institute of Research. There is no objection to its presentation/publication. The opinions or assertions contained herein are the private views of the author, and are not to be construed as official, or as reflecting true views of the Department of the Army or the Department of Defense. The investigators have adhered to the policies for protection of human subjects as prescribed in AR 70-25.

## Ethics

All participants provided written informed consent and samples were collected under approval the Ethical Review Committee for Research in Human Subjects, Ministry of Public Health (MoPH EC) and Walter Reed Army Institute of Research Institutional Review board under WRAIR #2902

## Author’s contributions

## References

1. Sudathip P, Kongkasuriyachai D, Stelmach R, Bisanzio D, Sine J, Sawang S, Kitchakarn S, Sintasath D, Reithinger R. The Investment Case for Malaria Elimination in Thailand: A Cost-Benefit Analysis. Am J Trop Med Hyg. 2019 Jun;100(6):1445–1453. doi: 10.4269/ajtmh.18-0897. PMID: 30994098; PMCID: PMC6553898.

2. Ashley EA, Dhorda M, Fairhurst RM, Amaratunga C, Lim P, Suon S, Sreng S, Anderson JM, Mao S, Sam B, Sopha C, Chuor CM, Nguon C, Sovannaroth S, Pukrittayakamee S, Jittamala P, Chotivanich K, Chutasmit K, Suchatsoonthorn C, Runcharoen R, Hien TT, Thuy-Nhien NT, Thanh NV, Phu NH, Htut Y, Han KT, Aye KH, Mokuolu OA, Olaosebikan RR, Folaranmi OO, Mayxay M, Khanthavong M, Hongvanthong B, Newton PN, Onyamboko MA, Fanello CI, Tshefu AK, Mishra N, Valecha N, Phyo AP, Nosten F, Yi P, Tripura R, Borrmann S, Bashraheil M, Peshu J, Faiz MA, Ghose A, Hossain MA, Samad R, Rahman MR, Hasan MM, Islam A, Miotto O, Amato R, MacInnis B, Stalker J, Kwiatkowski DP, Bozdech Z, Jeeyapant A, Cheah PY, Chalk J, Intharabut B, Silamut K, Lee SJ, Vihokhern B, Kunasol C, Imwong M, Tarning J, Smithuis F, Hlaing TM, Tun KM, Nwe ML, Barends M, Aung PP, van der Pluijm RW, Dhorda M, Imwong M, Woodrow CJ, Day NP, White NJ, Dondorp AM. Spread of artemisinin resistance in Plasmodium falciparum malaria. N Engl J Med. 2014 Jul 31;371(5):411–23. doi: 10.1056/NEJMoa1314981. PMID: 25075834; PMCID: PMC4085289.

3. World Health Organization. Artemisinin resistance and artemisinin-based combination therapy efficacy. World Health Organization. 2022. Available at: https://www.who.int/news-room/feature-stories/detail/artemisinin-resistance-and-artemisinin-based-combination-therapy-efficacy

4. Ménard D, Khim N, Beghain J, Adegnika AA, Shafiul-Alam M, Amodu O, Rahim-Awab G, Barnadas C, Berry A, Boum Y, Bustos MD, Cao J, Chen JH, Collet L, Cui L, Thakur GD, Dieye A, Djallé D, Dorkenoo MA, Eboumbou-Moukoko CE, Espino FE, Fandeur T, Ferreira-da-Cruz MF, Fola AA, Fuehrer HP, Hassan AM, Herrera M, Hongvanthong B, Houzé S, Ibrahim ML, Jahirul-Karim M, Jiang L, Kano S, Kassa M, Kengne P, Lanza M, Leang R, Leelawong M, Lin K, Mazarati JB, Mayxay M, Menard S, Morlais I, Muhindo-Mavoko H, Musset L, Na-Bangchang K, Nakazawa S, Namuigi P, Newton PN, Noedl H, Nour BY, Obare P, Ogouyèmi-Hounto A, Omar A, Pillai DR, Rogier C, Rosenthal PJ, Rossan RN, Same-Ekobo A, Sattabongkot J, Shi YP, Sutherland CJ, Swarthout T, Syafruddin D, Tahar R, Tang LH, Touré OA, Tsuyuoka R, Warsame M, Wini L, Zeyrek FY, Zinsou JF, Kwiatkowski D, Rogier C, Ringwald P, Ariey F, Mercereau-Puijalon O, Wongsrichanalai C, WorldWide Antimalarial Resistance Network Plasmodium falciparum K13 Genotype-Phenotype Study Group. A worldwide map of Plasmodium falciparum K13-propeller polymorphisms. N Engl J Med. 2016 Jun 23;374(25):2453–64. doi: 10.1056/NEJMoa1513137. PMID: 27332904; PMCID: PMC5034794.

5. Pholwat S, Liu J, Stroup S, Jacob ST, Banura P, Moore CC, Huang F, Laufer MK, Houpt E, Guler JL. The Malaria TaqMan Array Card Includes 87 Assays for Plasmodium falciparum Drug Resistance, Identification of Species, and Genotyping in a Single Reaction. Antimicrob Agents Chemother. 2017 Apr 24;61(5):e00110–17. doi: 10.1128/AAC.00110-17. PMID: 28264857; PMCID: PMC5404514.

6. Myo Thura Zaw, Tin Aung Hlaing, Aye Mya Thiri Kyaw, Cherry Wah Wah Myint, Jotshna Yonemori, Stuart D. Tyner, Nay Soe Maung, Kan Nagao. 2020. Prevalence of drug-resistant malaria in asymptomatic populations in myanmar: A nationwide study. The Journal of Infectious Diseases. DOI: 10.1093/infdis/jiz607

